# Autoantibodies to truncated GAD(96-585) antigen stratify risk of early insulin requirement in adult-onset diabetes

**DOI:** 10.1101/2023.11.30.23298881

**Authors:** Sian L. Grace, Kathleen M. Gillespie, Claire L. Williams, Vito Lampasona, Peter Achenbach, Ewan R. Pearson, Alistair J.K. Williams, Anna E. Long, Timothy J. McDonald, Angus G. Jones

**Author notes:** AEL, TJM and AGJ are joint senior authors. Corresponding author: Angus G. Jones. Address: University of Exeter Medical School, RILD Building, RD&E Hospital, Wonford, Barrack Road, Exeter, EX2 5DW.

## Abstract

**Objective:** We investigated whether further characterisation of full-length (f-) GADA responses could identify early insulin requirement in adult-onset diabetes.

**Research Design and Methods:** In 179 f-GADA positive participants diagnosed with type 2 diabetes, we assessed the association of truncated (t-)GADA positivity, f-GADA IgG subclasses, and f-GADA affinity with early insulin requirement (<5 years), type 1 diabetes genetic risk score (T1D GRS), and C-peptide. These characteristics were compared to f-GADA positive type 1 diabetes (n=141) and f-GADA negative type 2 diabetes (n=6420) cohorts.

**Results:** t-GADA positivity was lower in f-GADA positive without early insulin in comparison to f-GADA positive type 2 diabetes requiring insulin within 5 years, and type 1 diabetes (75% vs. 91% and 95% respectively, p<0.0001). t-GADA positivity (in those f-GADA positive) identified a group with a higher type 1 diabetes genetic susceptibility (mean T1D GRS 0.248 vs. 0.225, p=0.003), lower C-peptide (1156 pmol/L vs. 4289 pmol/L, p=1×10^-7^), and increased IA-2A positivity (23% vs. 6%, p=0.03). In survival analysis, t-GADA positivity was associated with early insulin requirement compared with those only positive for f-GADA, independently from age of diagnosis, f-GADA titre and duration of diabetes [adjusted HR 5.7 (95% CI 1.4, 23.5), p=0.017]. Early insulin requirement was not associated with an IgG1-restricted f-GADA response (p=0.81) or a high affinity f-GADA response (p=0.89).

**Conclusions:** The testing of t-GADA in f-GADA positive individuals with type 2 diabetes identifies those who have genetic and clinical characteristics comparable to type 1 diabetes and stratifies those at higher risk of early insulin requirement.

**Article Highlights:**

- Progression to insulin therapy is highly variable in adult-onset GADA positive diabetes.
- We further characterised GADA characteristics in adult-onset diabetes and assessed whether these are associated with early insulin requirement.
- Truncated GADA positivity was associated with a type 1 diabetes like phenotype and stratified risk of early insulin requirement. Those GADA positive who were negative for truncated GADA had the characteristics and progression of classical type 2 diabetes. Assessing full-length GADA IgG subclass and affinity did not further stratify risk of progression.
- t-GADA assessment remains underutilised in clinical practice, but could assist correct therapy allocation in adult-onset diabetes.

## Introduction

Autoantibodies to GAD are common in adults initially diagnosed and treated as type 2 diabetes, with prevalence varying from 2 to >10% depending on population and assay (1). This patient group, often described as having latent autoimmune diabetes (LADA), recently redefined by the WHO as slowly evolving immune-mediated diabetes (2; 3), is highly heterogeneous, varying from those with very rapid progression to insulin therapy and a type 1 diabetes like phenotype, to those with the clinical course and characteristics of type 2 diabetes.

Whether this heterogeneity is best explained by an intermediate form of autoimmune diabetes, or a mixture of autoimmune and non-autoimmune diabetes, due to the combination of imperfect islet autoantibody specificity and low prior likelihood of autoimmune diabetes (Bayes Theorem) in an adult population, or both, is a matter of debate (4-6). Approaches that improve specificity of GADA testing for identifying patients with type 1 diabetes would allow targeting of intensive monitoring, advice, and early insulin initiation to those most likely to benefit.

Developments in assay technology allow measurement of additional characteristics beyond full-length (f-)GADA titre, including epitope specificity, affinity, and IgG subclasses (7-9). The clinical utility of these GADA characteristics is unclear. Previous research in prediction has shown that GADA reactive to the n-terminally truncated GAD antigen (GAD96-585; t-GADA) are more disease-specific in first-degree relatives of patients with type 1 diabetes, whilst maintaining sensitivity and specificity in newly diagnosed cases (8; 10). Reactivity to t-GADA (11) and high f-GADA affinity (12) have been associated with risk of early insulin treatment in adult-onset diabetes, and increased IgG3 and IgG4 IgG subclasses have been associated with a slower rate of beta cell destruction in LADA (13).

We aimed to determine whether assessment of GADA truncated epitope specificity, affinity, and IgG subclasses within those with post diagnosis f-GADA positive type 2 diabetes, could improve the identification of patients with early insulin requirement and the C-peptide and genetic characteristics of type 1 diabetes.

## Research Design and Methods

### Study Cohorts

Participants included in this study were identified from 6 UK cohorts recruited from primary and secondary care and had uniform islet-autoantibody assessment (14): the Genetics of Diabetes Audit and Research Tayside Study (GoDarts) (15), Diabetes Alliance for Research in England (DARE) (16), Predicting Response to Incretin Based Agents in Type 2 Diabetes (PRIBA) (17), MRC MASTERMIND Progressors (18) and StartRight Studies (19; 20). A comparison cohort of participants with type 1 diabetes were also identified from DARE.

We assessed f-GADA characteristics in 179 f-GADA positive participants with a clinical diagnosis of type 2 diabetes after ≥age 18, and no insulin requirement within 6 months of diabetes (Supplemental Table 1). We compared islet-autoantibody, genetic and C-peptide characteristics with 6,420 participants with f-GADA negative type 2 diabetes (clinical diagnosis and >6 months to insulin), and 141 participants with type 1 diabetes (f-GADA positive, on insulin therapy from diagnosis, clinical diagnosis of type 1 diabetes). Characteristics for all cohorts are shown in Supplemental Tables 1-3.

### Assessment of HbA1c and Diabetes Progression (Time to Insulin)

Available HbA_1c_ at latest follow-up [median diabetes duration 11 years (range 7-15)] was obtained from electronic health care records for the GoDarts study (n=3,893) or was measured on a research sample in recruitment centers’ local laboratories (all are accredited NHS blood science laboratories) for the Exeter cohorts (PRIBA, MRC Progressors, StartRight, DARE; n=2,706).

For GoDARTS, time to insulin was defined from electronic prescription records. For Exeter cohorts (DARE, PRIBA, MRC MASTERMIND Progressors), insulin treatment, date of commencing insulin, and date of diagnosis were self-reported at a single visit. For StartRight, insulin treatment, date of commencing insulin, and date of diagnosis were self-reported at three visits, within 12 months of diagnosis, and at approximately 1 and 2 years later (21).

### Laboratory Measurement of GADA to full-length GAD65(1-585)

Analysis of f-GADA was conducted at The Academic Department of Blood Sciences, Royal Devon and Exeter Hospital using the RSR Limited ELISA (Cardiff, U.K.) on the Dynex DS2 ELISA Robot (Dynex Technologics, Worthing, U.K.). The cut-off for positivity was ≥11 World Health Organisation (WHO) units/mL, based on the 97.5^th^ centile of 1,559 control participants without diabetes (22). In the 2020 International Islet Autoantibody Standardization Program (IASP) the assay specificity and adjusted sensitivity at 95% specificity (AS95) were 98.9% and 86%, respectively.

### Assessment of GADA characteristics

Of 6,599 participants, 179 (2.7%) were f-GADA positive with sera available for further characterisation. These, and 141 f-GADA positive patients with type 1 diabetes, underwent further analysis to explore autoantibody characteristics: t-GADA epitope specificity, f-GADA affinity, and f-GADA IgG subclasses.

#### Measurement of GADA epitope specificity to truncated GAD65(96-585)

t-GADA epitope specificity was determined by a luciferase immunoprecipitation system (LIPS) assay using nanoluciferase-tagged GAD65/67 kDa isoform of GAD antigen, with the n-terminal 95aa truncated (Nluc-GAD65(96-585)), as described previously (23). Diabetic kidney units/ml (DK U/ml) were calculated using a logarithmic standard curve and the threshold of positivity was ≥10.7 DK units/ml (based on the 97.5th centile of 221 healthy schoolchildren). In the IASP2020 workshop, the specificity and AS95 for this assay were 100% and 86%, respectively.

#### Measurement of GADA IgG subclass response to full-length GAD65(1-585)

Determination of IgG subclasses to f-GADA was based on a previously published approach with modifications (9; 24), described in detail in ESM Supplemental Methods.

Due to serum availability, a sub-cohort of f-GADA positive samples were selected for subclass analysis, ensuring an equal number of samples with f-GADA positive type 2 diabetes with and without progression to insulin within 5 years, and f-GADA positive type 1 diabetes. Samples from each cohort studied were run simultaneously in each assay, and where possible, were matched for f-GADA titre and affinity (closest available) across comparison groups.

#### Measurement of GADA Affinity to full-length GAD65(1-585)

Affinity of f-GADA was measured by competitive binding experiments based on the approach developed by Mayr *et al* (7), described in detail in ESM Supplemental Methods, using increasing concentrations of unlabelled human GAD65.

The calculation of *K*_d_ values was limited to samples with IC_50_ values greater than the concentration of labelled GAD65 (1.88×10^-10^ mol/l). For samples with an IC_50_ <1.88×10^-10^ mol/L, the f-GADA affinity of the sample was set at *K*_d_ >8×10^11^ l/mol. A negative QC sample (healthy adult) and a positive QC sample [f-GADA positive relative without diabetes, (38% CV)] was run alongside samples in each assay.

### Assessment of IA-2A & ZnT8A Positivity

IA-2A & ZnT8A assessment was undertaken in all samples from DARE, StartRight, MRC Progressors, PRIBA and GoDarts (ZnT8A only) was conducted on the same serum sample as the f-GADA assessment at The Academic Department of Blood Sciences, Royal Devon and Exeter Hospital using the RSR Limited ELISAs on the Dynex DS2 ELISA Robot. The cut-off for IA-2A positivity was ≥7.5 units/mL, based on the 97.5^th^ centile of 1,559 control participants without diabetes (22). In the IASP2020 workshop, the specificity and AS95 was 98.9% and 72%, respectively. ZnT8A positivity was ≥65 WHO units/mL for those aged <30 years and ≥10 WHO units/mL for those aged ≥30 years, based on the 97.5^th^ centile of 1,559 control participants without diabetes (26). In the IASP2020 workshop, the assay specificity and AS95 were 98.9% and 74%, respectively. As IA-2A was not measured in GoDarts, to ensure complete data for the f-GADA positives, IA-2A was remeasured on all 179 and the f-GADA positives with type 1 diabetes, using a LIPS assay, as described above, but using a Nluc-tagged antigen specific to the intracytoplasmic (aa606-979) region of islet antigen-2 (IA-2ic) kindly provided by Vito Lampasona (Milan, Italy). The threshold of positivity was ≥0.3 DK U/ml (based on the 98th centile of 112 school children). In the IASP2020 workshop, the specificity and AS95 for this assay were 100% and 78%, respectively.

### Additional laboratory analysis (C-peptide and type 1 diabetes genetic risk score)

Plasma C-peptide was measured by electrochemiluminescence immunoassay (intra-assay CV, 3.3%; inter-assay CV, 4.5%) on a Roche Diagnostics (Mannheim, Germany) E170 analyser by the Blood Sciences Department at the Royal Devon and Exeter NHS Foundation Trust (Exeter, U.K.)

We generated weighted T1D-GRS from 30 common type 1 diabetes genetic variants [single nucleotide polymorphisms (SNPs)] for HLA and non-HLA loci as previously described (14; 27).

### Statistical Analysis

We compared proportions of the following GADA characteristics [t-GADA status (positive vs. negative), IgG subclass response (IgG1-restricted vs. IgG-unrestricted) and affinity (high vs. moderate/low affinity)] between f-GADA positive clinically diagnosed type 2 diabetes with and without early insulin treatment (<5 years) and f-GADA positive type 1 diabetes using the Pearson Chi2 test. We then assessed whether each characteristic was associated with clinical and biochemical participants’ characteristics within all those with f-GADA positive type 2 diabetes using Pearson chi-squared tests for proportions of categorical variables [IA-2A & ZnT8A positivity and early insulin requirement (<5 years)] and t-tests for continuous variables (C-peptide, T1D-GRS, f-GADA titre and age-at-diagnosis).

We assessed the relationship between GADA characteristics and progression to insulin (censored at 5 years) using cox proportional hazard models (after confirming model assumptions) in univariable and multivariable models, with adjustment for f-GADA titre, duration of diabetes at f-GADA test, and age-at-diagnosis). For f-GADA affinity, we also assessed whether there was an association between higher affinity and IgG1-restricted responses and progression to insulin therapy independent of t-GADA specificity in addition to the above co-variates. All statistical analysis was carried out using Stata/SE 16.0 (StataCorp, College Station, TX) unless otherwise stated and graphed using GraphPad Prism3.

### Data and Resource Availability

The StartRight dataset generated during and/or analysed in the current study is available from the corresponding author upon reasonable request. Data pertaining to the other Exeter studies (DARE, PRIBA and MRX Progressors) can be accessed via application to the Penninsula Research Bank; and for GoDARTs via application to the GoDARTs study committee.

## Results

In those with f-GADA positive type 2 diabetes (n=179), median follow up was 12 years, with f-GADA assessment at a median of 4.9 years diabetes duration; 35% (n=63) of participants had progressed to insulin ≤5 years. In the comparison cohorts; those with f-GADA negative type 2 diabetes (n=6,420) and f-GADA positive type 1 diabetes (n=141); median follow-up was 11 and 15 years, and f-GADA assessment was at a median 5.6 and 16 years diabetes duration, respectfully.

### Participants with positive GADA for a truncated epitope have enrichment for genetic and clinical characteristics associated with type 1 diabetes

Positivity for t-GADA was similar between individuals with f-GADA positive type 1 diabetes and those with f-GADA positive type 2 diabetes requiring early insulin (≤5 years) 95% (95% CI 90, 98) vs. 97% (95% CI 89, 100) respectively, p=0.57). In contrast, the proportion of those t-GADA positivity without early insulin requirement was significantly lower [72% (95% CI 63, 80)] than individuals with early insulin requirement (p=7×10^-5^) and the type 1 diabetes cohort (p=4×10^-7^)) (Figure 1A). t-GADA positivity identified a group diagnosed younger [mean 55 years (95% CI 52, 57) vs. 62 years (95% CI 58, 66), p=0.002], with a higher T1D-GRS [mean 0.248 (95% CI 0.241, 0.254) vs. 0.225 (95% CI 0.213, 0.237), p=0.003], lower c-peptide levels [mean 1155 pmol/L (95% CI 918, 1393) vs. 4289 pmol/L (95% CI 845, 7732), p=1×10^-7^ at a median duration of 12 years at C-peptide testing] and increased positivity for IA-2A [23% (95% CI 17, 31) vs. 6% (95% CI 0.7, 19.7), p=0.022] and ZnT8A [21% (95% CI 14, 28) vs. 0% (95% CI 0, 10), p=0.004] than those positive for f-GADA but t-GADA negative (Table 1).

**Figure 1:**
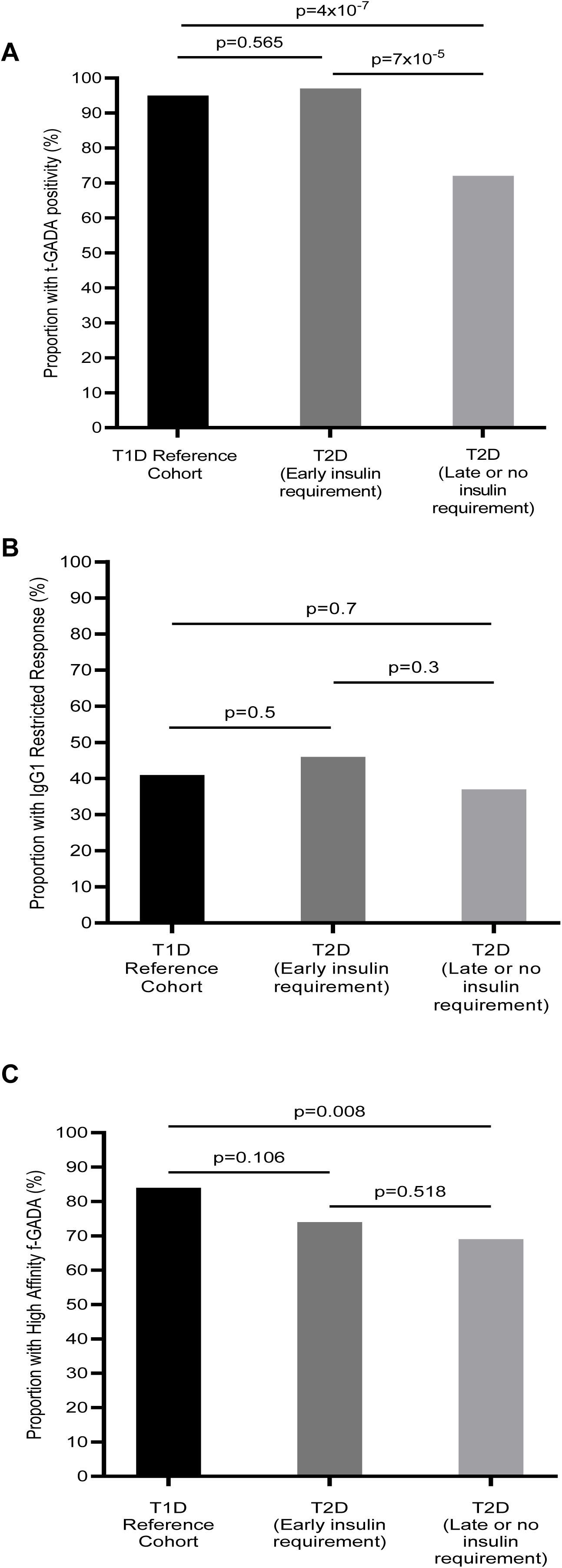
Proportions of individuals with t-GADA (A), IgG1-restricted f-GADA response (B) and high affinity f-GADA response (C). T1D; Type 1 diabetes. T2D; type 2 diabetes. t-GADA; truncated GAD(96-585) autoantibody. f-GADA; full length GAD(1-585) autoantibody.

**Table 1:**
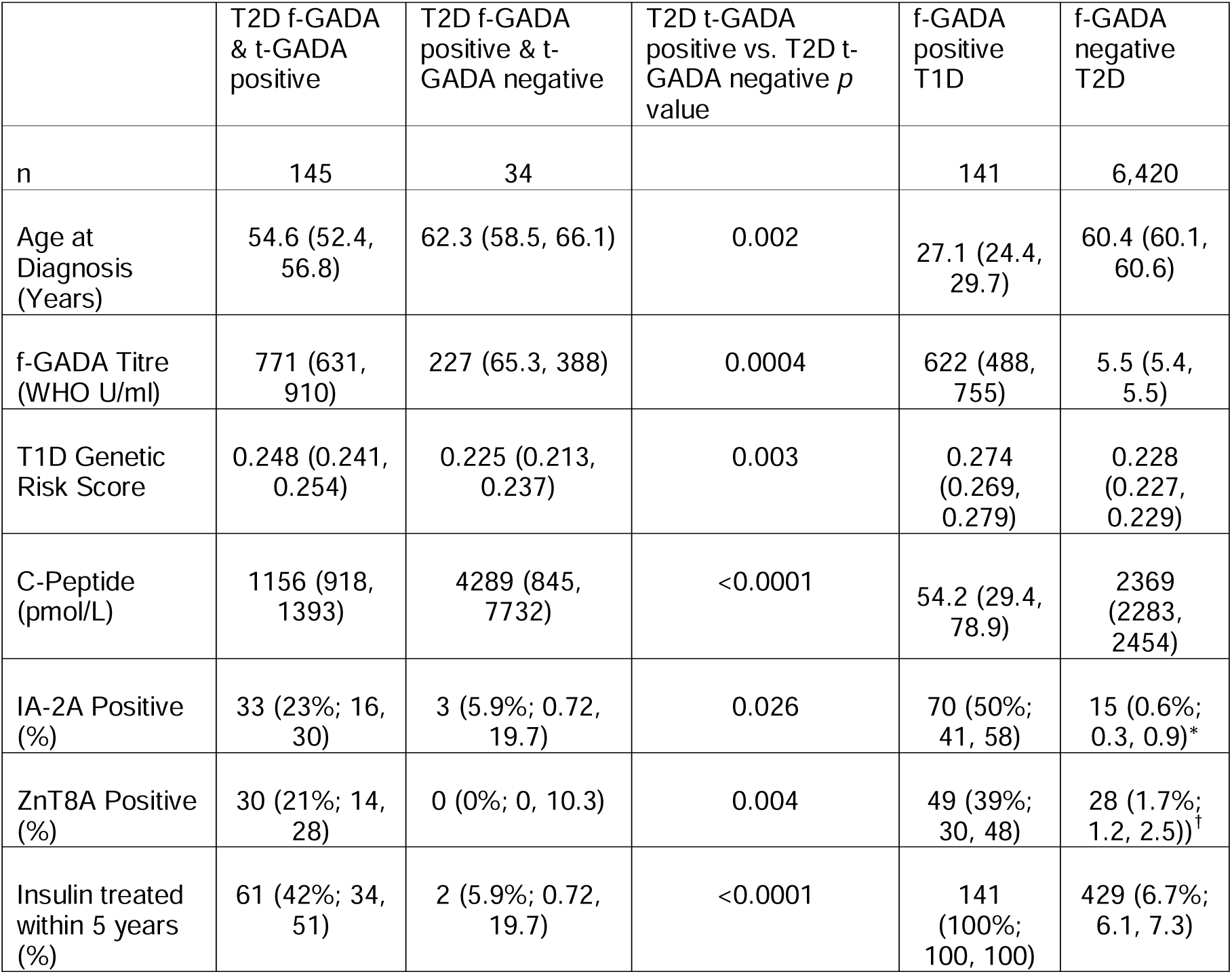
Diabetes characteristics comparison between those positive and negative for t-GADA in those f-GADA positive. Data displayed as n(%; 95% CI) or mean (95% CI). *Out of 2,607 tested. ^†^Out of 1,615 tested. T1D; Type 1 Diabetes. T2D; Type 2 Diabetes. t-GADA; truncated GAD(96-585) autoantibody. f-GADA; full length GAD(1-585) autoantibody. IA-2A; islet antigen-2 autoantibody. ZnT8A; zinc transporter 8 autoantibody.

### Truncated GADA epitope positivity is independently associated with increased risk of early insulin therapy

In survival analysis t-GADA positivity (in those f-GADA positive) identified participants at higher risk of early progression to insulin compared to those f-GADA positive & t-GADA negative [HR 8.4 (95% CI 2.1, 34.4), p=0.003] (Table 2; Figure 2A). The association between t-GADA positivity (in those f-GADA positive) and early insulin requirement persisted after adjustment for age-at-diagnosis, f-GADA titre and duration at GAD testing [adjusted HR 5.7 (95% CI 1.4, 23.5), p=0.017] compared to those f-GADA positive and t-GADA negative (Table 2). Those positive for f-GADA but negative for t-GADA had similar risk of progression to early insulin requirement compared to those with f-GADA negative type 2 diabetes [HR 0.93 (95% CI 0.23, 3.72), p=0.9]. This was similar after adjustment for age-at-diagnosis, f-GADA titre and diabetes duration at GAD testing [adjusted HR 0.98 (95% CI 0.24, 3.95), p=0.98] (Supplemental Table 4).

**Figure 2:**
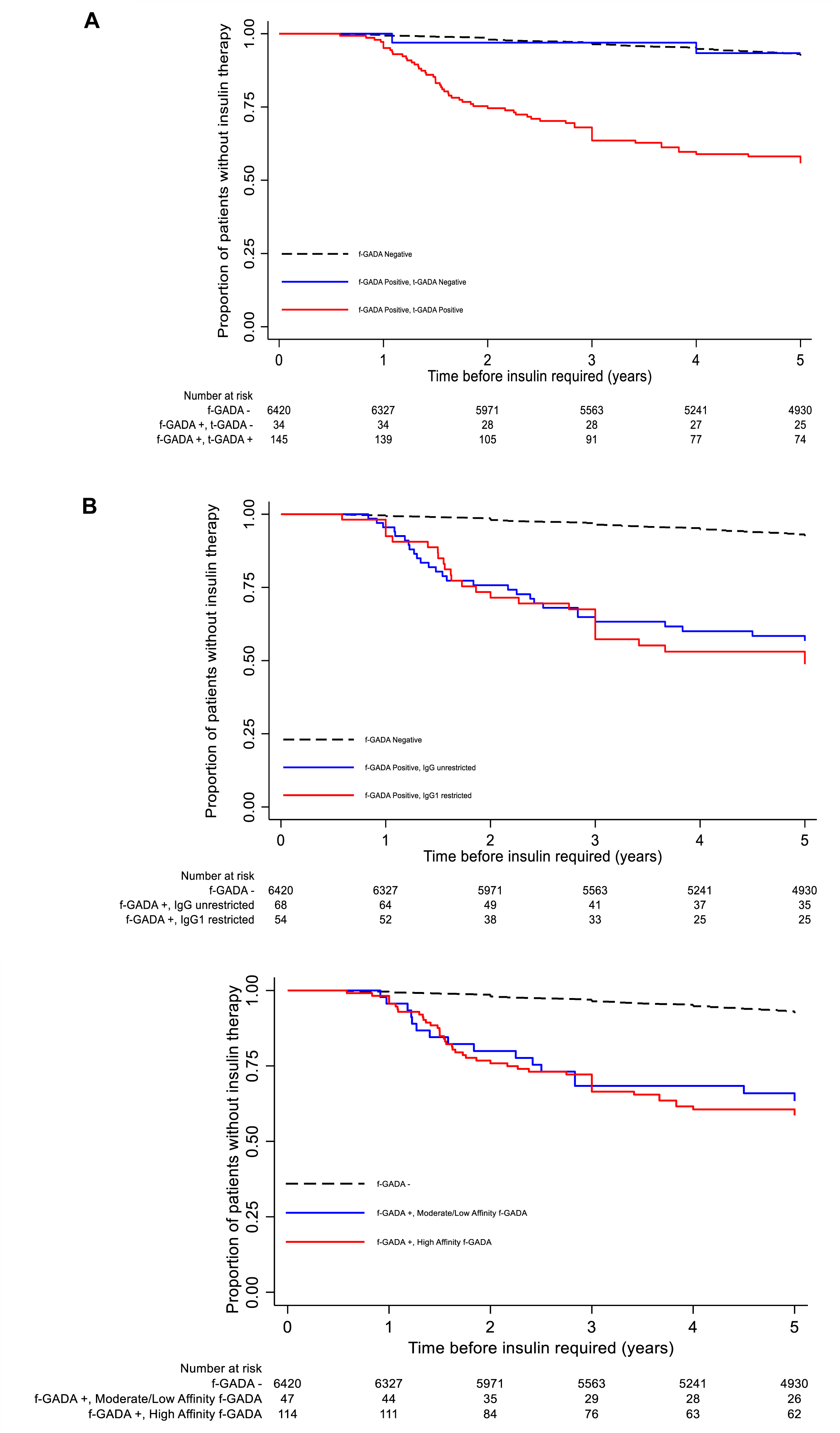
Kaplan-Meier plots of probability of requiring insulin therapy during 5-year follow-up, in those clinically diagnosed with type 2 diabetes. A) Stratified by risk group of f-GADA and t-GADA positivity. Solid lines represent f-GADA positive groups and dashed lines represent f-GADA negative group. Blue line indicates t-GADA negative and red line is t-GADA positive. B) Stratified by risk group of f-GADA positivity and subclass. Solid lines represent f-GADA positive groups and dashed line represent f-GADA negative group. Blue line indicates IgG unrestricted response and red line is IgG1 restricted response. C) Stratified by risk group of f-GADA positivity and affinity. Solid lines represent f-GADA positive groups and dashed line represent f-GADA negative group. Blue line indicates a moderate/low affinity f-GADA response and red line is a high affinity f-GADA response. +, positive. -, negative.

**Table 2:** Hazard Ratios from Cox proportional regression models, stratified by t-GADA status in those f-GADA positive, (unadjusted and adjusted) for time to insulin censored at 5 years. t-GADA; truncated GAD(96-585) autoantibody. f-GADA; full length GAD(1-585) autoantibody.

| **Survival analysis t-GADA pos vs t-GADA neg (In those f-GADA positive)** | **Unadjusted Model** |  | **Adjusted Model** |  |
| --- | --- | --- | --- | --- |
|  | **<u>Hazard ratio (95% CI)</u>** | **<u>p value</u>** | **<u>Hazard ratio (95% CI)</u>** | **<u>p value</u>** |
| t-GADA negative (reference) | 1 |  | 1 |  |
| t-GADA positive (vs. reference) | 8.4 (2.05, 34.4) | 0.003 | 5.7 (1.4, 23.5) | 0.017 |
| Age of Diagnosis (per 1 year increase) |  |  | 0.94 (0.92, 0.96) | <0.001 |
| f-GADA Titre (per 100 unit increase) |  |  | 1.01 (0.98, 1.04) | 0.536 |
| Duration of Diabetes at f-GADA testing (per 1 year increase) |  |  | 0.88 (0.83, 0.94) | <0.001 |

### Full-length GADA IgG subclasses do not identify those at risk of early insulin therapy

The prevalence of each f-GADA IgG subclass was similar between f-GADA positive type 2 diabetes participants with and without early insulin requirement and those with f-GADA positive type 1 diabetes (p>0.07 for all comparisons, Supplemental Table 5). The rank order of frequencies of IgG subclasses was the same between those with type 2 diabetes and early insulin requirement and those without early insulin requirement (IgG1>IgG3>IgG2>IgG4). In the f-GADA positive with type 1 diabetes reference group, the rank order of frequencies of IgG subclasses was IgG1>IgG3>IgG4>IgG2. f-GADA IgG subclasses were unable to be detected in 13 (6%) of the subset tested. As IgG1 was the most common IgG subclass in all three cohorts, we split the cohort into two response categories for further analysis: IgG1 only (IgG1-restricted) vs. IgG1 + other IgG subclasses (IgG-unrestricted). The proportion of those with an IgG1-restricted response was similar between those with type 2 diabetes and early insulin requirement vs. those without early insulin requirement [42% (95% CI 29, 57) vs. 39% (95% CI 28, 52), p=0.7]. The proportion of those with an IgG1-restricted response in the f-GADA positive type 1 diabetes group was similar [40% (95% CI 29, 53), p vs. other subgroups >0.8] (Figure 1B). IgG subclass response was not associated with clinical characteristics (age-at-diagnosis, T1D-GRS, c-peptide levels, and IA-2A & ZnT8A positivity), but those with an IgG1-restricted response had lower levels of f-GADA than those with an IgG-unrestricted response (mean 468 WHO U/ml (95% CI 283, 652) vs. 1130 WHO U/ml (95% CI 918, 1342), p<0.0001 (Supplemental Table 6).

In survival analysis, an IgG1-restricted response did not identify those at risk of early insulin requirement in those that were f-GADA positive [HR 1.07 (95% CI 0.62, 1.9), p=0.8] (Figure 2B). This was still the case when the model was adjusted for age-at-diagnosis and duration of diabetes [HR 1.02 (95% CI 0.58, 1.8), p=0.9] (Supplemental Table 7). The presence of each individual IgG subclass was not associated with progression to insulin in survival analysis (Supplemental Table 8).

### The proportion of high affinity full-length GADA was lower in those with type 2 diabetes

The affinities of f-GADA detected ranged from 7.57×10^6^ to >8 x10^11^ l/mol across all groups (type 2 diabetes with early insulin requirement 3.94 x10^7^ to >8 x10^11^ l/mol, type 2 diabetes with no/later insulin requirement 7.57 x10^6^ to >8 x10^11^ l/mol, f-GADA positive type 1 diabetes 3.76×10^7^ to >8 x10^11^ l/mol). For categorial analysis, affinities were split into high (≥1×10^9^ l/mol) and moderate/low affinity groups (<1×10^9^ l/mol) in line with previous publications (7; 12; 28). The proportion of those with high affinity f-GADA was similar between those with type 2 diabetes with and without early insulin requirement [74% (95% CI 61, 84) vs. 69% (95% CI 59, 78), p=0.5]. Those with f-GADA positive type 1 diabetes had a higher proportion of those with high affinity f-GADA [84% (95% CI 76, 89)] compared to those with early insulin requirement (p=0.1) and without (p=0.008) (Figure 1C). There were no differences in age-at-diagnosis, c-peptide levels, and IA-2A & ZnT8A positivity between those with high and moderate/low affinity (Supplemental Table 9). However, those with high affinity f-GADA had lower f-GADA titres [mean 546 WHO U/ml (95% CI 409, 683) vs. mean 1167 WHO U/ml (95% CI 902, 1432), p=1×10^-5^] and higher T1D GRS [mean 0.249 (95% CI 0.242, 0.256) vs. mean 0.232 (96% CI 0.219, 0.244), p=0.01] than those with moderate/low affinity f-GADA.

Stratification by f-GADA affinity category in those f-GADA positive did not stratify risk of progression to insulin therapy [HR 1.13 (95% CI 0.64, 2.01), p=0.66] (Figure 2C). Again, this was still the case when the model was adjusted for age-at-diagnosis, f-GADA titre, and duration of diabetes at f-GADA testing [HR 1.17 (95% CI 0.63, 2.17), p=0.62] (Supplemental table 10). f-GADA affinity did not further stratify early insulin requirement in those found to be t-GADA positive (Supplemental Figure 1).

## Conclusions

Our study shows that in individuals with f-GADA positive type 2 diabetes, testing for t-GADA identified those with a more type 1 diabetes like phenotype (diagnosed younger, increased proportion positive for multiple islet autoantibodies, increased type 1 diabetes genetic susceptibility, and lower c-peptide levels), and stratifies risk of early insulin requirement. Whilst t-GADA positivity is strongly associated with early insulin requirement, participants positive for f-GADA but negative for t-GADA had similar risk of early insulin requirement to f-GADA negative type 2 diabetes. In contrast, assessment of f-GADA affinity and IgG subclass response did not further stratify risk of early insulin requirement over and above f-GADA testing, and (with the exception of affinity and T1D GRS) were not associated with other characteristics of type 1 diabetes.

To our knowledge this is the first study to show that t-GADA identified those that are at risk of early insulin requirement independently of f-GADA titre, duration of diabetes at GADA assessment, and age-at-diagnosis. This is the first study to assess the relationship between t-GADA epitope, f-GADA affinity and IgG subclass response (in the same cohort) and early insulin treatment using survival analysis, and the first to compare these characteristics in a large cohort with f-GADA positivity assessed using a highly specific, clinically available assay. Our finding of positivity for t-GADA, identifying those at higher risk of progressing to insulin therapy with a more type 1 diabetes-like phenotype is consistent with Achenbach *et al* (11), however the clinical utility of truncated GADA in predicting early insulin in survival analysis (in a cohort with longer follow-up), and the association with higher type 1 diabetes genetic susceptibility and lower c-peptide were not previously described.

Like Hillman *et al* (13), we found the presence of all IgG subclasses of f-GADA present in our adult-onset type 2 diabetes cohort, with similar proportions observed for IgG1, IgG2, and IgG4. However, we also observed a higher proportion of IgG3 in our type 2 diabetes cohort (50% vs. 10% observed by Hillman *et al.*), which could be due to improvements in the clone used for the anti-human IgG3 antibody and the previous one used by previous publications being discontinued. We observed the presence of IgG4 in our type 1 diabetes reference cohort (21% had f-GADA IgG4) which Hillman *et al* did not previously observe. The difference in the presence of IgG4 subclass in our type 1 diabetes reference cohort may be due to the differences in duration of diabetes at analysis, with Hillman *et al* testing samples within days of diagnosis while we tested at a median of 16 years post diagnosis. The presence of IgG4 subclasses in our study could be due to prolonged stimulation and maturation of B cells and GADA responses remaining highly prevalent post-diagnosis (29).

We have shown that higher affinity f-GADA do not identify those at a higher risk of early insulin requirement or those with a more type 1 diabetes-like phenotype, in contrast to previous research (12; 30). This may be due to the wide variation in affinities found in the studies, differences in f-GADA screening and affinity assay format, differences in duration of diabetes at testing or to differences in what is described as higher or lower affinity. However, our study did have substantially larger numbers compared to Krause *et al* (n=163 vs. n=47) and a longer duration of follow up.

A strength of our study is the size and detailed follow-up data of the initial adult-onset cohort with type 2 diabetes (>6,000) screened for f-GADA in one laboratory, using a highly robust and specific bridge ELISA assay and a positivity threshold based on a large control population. This is a highly unique cohort as we had follow-up C-peptide data from diagnosis as well as T1D GRS data. We were also able to apply a series of well-developed strategies and high-quality tests to examine in detail the characteristics of GADA in this well-defined cohort and compare them to a cohort with f-GADA positive type 1 diabetes. To improve upon the clinical ELISA assay (used as the f-GADA screen in this study), future work could consider trying to incorporate the n-terminally truncated assay into the plate format, as whilst the t-GADA LIPS assay can be used for screening in a research setting, it is not set up on an automated platform.

A caveat of our research is that t-GADA testing was applied only to participants positive for a f-GADA assay due to time, sample availability, and cost constraints. As we have not tested those negative for the full length assay, our results can only currently be applied to those that have previously tested f-GADA positive, and findings for the whole cohort (including f-GADA negative participants) should be treated with caution, as it is possible that false positive results for t-GADA could occur in the >6000 of our cohort not tested for t-GADA which could blunt the diagnostic accuracy and hazard ratios of t-GADA testing reported for the whole cohort in this study. Previously, Williams *et al*, identified 1% of those that previously tested f-GADA negative to be t-GADA positive (8). Such low rates of t-GADA positivity in those who are f-GADA negative could lend support to the assumption that t-GADA is likely to have high specificity when applied to a whole population.

The f-GADA characterisation assays used in this study were conducted in different assay formats to the initial screening assay. Our original f-GADA screen was conducted using the RSR ELISA assay, whilst the f-GADA characteristics were conducted using liquid-phase radiobinding and LIPS assays. Differences between the ELISA and liquid-phase assays have been reported to impact on specificity and sensitivity (31; 32). IgG subclasses, epitope, and affinity characteristics are unable to be assessed via RSR bridging ELISA assays at this time. As the RSR bridging ELISA is the most commercially and clinically used f-GADA assay (and the assay type with highest overall performance in the international autoantibody standardisation program (33)), this allows us to compare the t-GADA LIPS assay with a highly specific and currently used assay in the clinical setting. A further limitation is that f-GADA positivity was assessed in all patients with type 2 diabetes at a median 5.6 years after diabetes diagnosis, f-GADA prevalence is likely to be lower than at diagnosis, although in adult-onset diabetes differences at this duration are modest (14; 34).

Diagnosing autoimmune diabetes in later life is an important and challenging clinical problem, and full-length GAD assays are unlikely to be sufficiently specific to confirm autoimmune diabetes in the setting of those diagnosed initially as type 2 diabetes (4; 6; 35). Therefore, approaches that improve islet autoantibody test specificity are needed to improve identification of autoimmune diabetes in adults (4). Our findings suggest that assays using t-GADA may have improved performance for identification of patients with early progression and the phenotype of type 1 diabetes, potentially improving identification of these patients in clinical practice and research. Thus, providing more support to the argument that t-GADA testing can replace or add to f-GADA testing in a clinical setting. Assays for f-GADA affinity or IgG subclass are more expensive, requiring specialist reagents and techniques and do not lend themselves readily to testing in a clinical setting, and our finding that they are unlikely to improve identification of this group above testing for t-GADA suggests they are unlikely to have clinical utility for this purpose.

In conclusion, the testing of t-GADA in f-GADA positive individuals with type 2 diabetes identifies those who have genetic and clinical characteristics comparable to type 1 diabetes and stratifies those at higher risk of early insulin requirement.

## Funding and Assistance

This study was supported by a grant from The NovoNordisk UK Research Foundation. SLG was funded by an E3 PhD Studentship. AEL is jointly funded as a Diabetes UK & JDRF RD Lawrence Fellow (18/0005778 and 3-APF-2018-591-A-N). CLW was funded by Diabetes UK (16/0005556 and 21/0006332). AGJ was supported for this work by an NIHR Clinician Scientist award (CS-2015-15-018). TJM is an NIHR Senior Clinical Senior Lecturer. ERP is a Wellcome Trust New Investigator (102820/Z/13/Z).

StartRight study is funded by Diabetes UK (17/0005624) and the UK National Institute of Health and Social Care Research (CS-2015-15-018). GADA assessment in GoDARTS was funded by EU Innovative Medicines Initiative 115317 (DIRECT), resources of which are composed of financial contributions from the European Union’s Seventh Framework Programme (FP7/2007-2013) and European Federation of Pharmaceutical Industries and Associations companies in kind contribution. The DARE study was funded by the Wellcome Trust and supported by the Exeter NIHR Clinical Research Facility. The MASTERMIND study was funded by the U.K. Medical Research Council (MR/N00633X/) and supported by the Exeter NIHR Clinical Research Facility. The PRIBA study was funded by the National Institute for Health Research (NIHR; U.K.; DRF-2010-03-72) and supported by the Exeter NIHR Clinical Research Facility. Genotyping for generation of the type 1 genetic risk score was supported by the European Foundation for the Study of Diabetes (2016 Rising Star Fellowship). This study was supported by the National Institute for Health and Care Research Exeter Biomedical Research Centre and National Institute for Health and Care Research Exeter Clinical Research Facility. The views expressed are those of the authors and not necessarily those of the NIHR or the Department of Health and Social Care.

## Acknowledgement of co-authors and contributions to paper

The authors thank Mrs Olivia Pearce, University of Bristol, for assisting in the laboratory analysis with the GADA characterisation and IA-2A LIPS assays. Mrs Pearce received no financial support for her participation. This work was conducted in collaboration with the UK T1D Research Consortium.

KMG, AJKW, TJM and AGJ initially designed the study. ERP, TJM and AGJ created and managed the research studies. SLG, AEL and CLW researched the GADA characteristic data using specialist assays created, developed and optimised by CLW, AJKW, VL and PA. SLG, AEL, KMG, TJM and AGJ analysed the data with advice from CLW and PA. SLG wrote the first draft of the manuscript with AEL, KMG, TJM and AGJ. All authors provided helpful discussion and reviewed and edited the manuscript. AGJ is the guarantor of this work and, as such, had full access to all the data in the study and takes responsibility for the integrity of the data and the accuracy of the data analysis.

## Prior Presentation

This data has been presented in part at the following conferences:

**Grace S.L.,** Long A.E., Gillespie K.M., Williams A.J.K., Lampasona V., Achenbach P., Pearson E.R., McDonald T.J, Jones A.G. (2022) “Glutamate decarboxylase islet autoantibody characteristics can stratify those at risk of rapid insulin progression in adult-onset type 2 diabetes (Oral Presentation) September 2022. *EASD*. Stockholm, Sweden.

**Grace S.L.,** Long A.E., Gillespie K.M., Williams A.J.K., Lampasona V., Achenbach P., Pearson E.R., McDonald T.J, Jones A.G. (2022) “Rapid insulin progression in glutamate decarboxylase autoantibody positive adult-onset diabetes can be better predicted by detecting autoantibodies to n-terminally truncated GAD(96-585)” (Poster Presentation – Shortlisted for Clinical Science Poster Award) *The Diabetes UK Professional Conference*. March 2022. Online

**Grace S.L.,** Long A.E., Gillespie K.M., Williams A.J.K., Lampasona V., Achenbach P., Pearson E.R., McDonald T.J, Jones A.G. (2022) “Autoantibodies specific to the n-terminally truncated GAD(96-585) can stratify rapid progressors to insulin therapy in adult-onset GAD(1-585) positive diabetes” (Poster Presentation) *Immunology of Diabetes Society Congress*. November 2021. Online

## References

1. Pieralice S, Pozzilli P. Latent Autoimmune Diabetes in Adults: A Review on Clinical Implications and Management. Diabetes Metab J 2018;42:451–464

2. WHO Diabetes Classification Guidelines 2019 [article online], 2019. Available from https://www.who.int/publications/i/item/classification-of-diabetes-mellitus. Accessed 10th October 2022

3. Buzzetti R, Zampetti S, Maddaloni E. Adult-onset autoimmune diabetes: current knowledge and implications for management. Nat Rev Endocrinol 2017;13:674–686

4. Jones AG, McDonald TJ, Shields BM, Hagopian W, Hattersley AT. Latent Autoimmune Diabetes of Adults (LADA) Is Likely to Represent a Mixed Population of Autoimmune (Type 1) and Nonautoimmune (Type 2) Diabetes. Diabetes Care 2021;44:1243–1251

5. Buzzetti R, Tuomi T, Mauricio D, Pietropaolo M, Zhou Z, Pozzilli P, Leslie RD. Management of Latent Autoimmune Diabetes in Adults: A Consensus Statement From an International Expert Panel. Diabetes 2020;69:2037

6. Steck AK, Eisenbarth GS. Genetic similarities between latent autoimmune diabetes and type 1 and type 2 diabetes. Diabetes 2008;57:1160–1162

7. Mayr A, Schlosser M, Grober N, Kenk H, Ziegler AG, Bonifacio E, Achenbach P. GAD Autoantibody Affinity and Epitope Specificity Identify Distinct Immunization Profiles in Children at Risk for Type 1 Diabetes. Diabetes 2007;56:1527

8. Williams AJ, Lampasona V, Wyatt R, Brigatti C, Gillespie KM, Bingley PJ, Achenbach P. Reactivity to N-Terminally Truncated GAD65(96-585) Identifies GAD Autoantibodies That Are More Closely Associated With Diabetes Progression in Relatives of Patients With Type 1 Diabetes. Diabetes 2015;64:3247–3252

9. Bonifacio E, Scirpoli M, Kredel K, Füchtenbusch M, Ziegler A-G. Early Autoantibody Responses in Prediabetes Are IgG1 Dominated and Suggest Antigen-Specific Regulation. The Journal of Immunology 1999;163:525

10. Pöllänen PM, Härkönen T, Ilonen J, Toppari J, Veijola R, Siljander H, Knip M. Autoantibodies to N-terminally Truncated GAD65(96-585): HLA Associations and Predictive Value for Type 1 Diabetes. The Journal of Clinical Endocrinology & Metabolism 2022;107:e935–e946

11. Achenbach P, Hawa MI, Krause S, Lampasona V, Jerram ST, Williams AJK, Bonifacio E, Ziegler AG, Leslie RD, on behalf of the Action Lc. Autoantibodies to N-terminally truncated GAD improve clinical phenotyping of individuals with adult-onset diabetes: Action LADA 12. Diabetologia 2018;61:1644-1649

12. Krause S, Landherr U, Agardh CD, Hausmann S, Link K, Hansen JM, Lynch KF, Powell M, Furmaniak J, Rees-Smith B, Bonifacio E, Ziegler AG, Lernmark A, Achenbach P. GAD autoantibody affinity in adult patients with latent autoimmune diabetes, the study participants of a GAD65 vaccination trial. Diabetes Care 2014;37:1675–1680

13. Hillman M, Törn C, Thorgeirsson H, Landin-Olsson M. IgG4-subclass of glutamic acid decarboxylase antibody is more frequent in latent autoimmune diabetes in adults than in type 1 diabetes. Diabetologia 2004;47:1984–1989

14. Grubb AL, McDonald TJ, Rutters F, Donnelly LA, Hattersley AT, Oram RA, Palmer CNA, van der Heijden AA, Carr F, Elders PJM, Weedon MN, Slieker RC, t Hart LM, Pearson ER, Shields BM, Jones AG. A Type 1 Diabetes Genetic Risk Score Can Identify Patients With GAD65 Autoantibody-Positive Type 2 Diabetes Who Rapidly Progress to Insulin Therapy. Diabetes Care 2019;42:208-214

15. Hébert HL, Shepherd B, Milburn K, Veluchamy A, Meng W, Carr F, Donnelly LA, Tavendale R, Leese G, Colhoun HM, Dow E, Morris AD, Doney AS, Lang CC, Pearson ER, Smith BH, Palmer CNA. Cohort Profile: Genetics of Diabetes Audit and Research in Tayside Scotland (GoDARTS). Int J Epidemiol 2018;47:380–381j

16. Diabetes Alliance For Research In England (DARE) [article online], 2017. Available from https://www.diabetesgenes.org/current-research/dare/#:~:text=The%20DARE%20Study%20is%20a,the%20Exeter%20region%20and%20nationwide.

17. Jones AG, McDonald TJ, Shields BM, Hill AV, Hyde CJ, Knight BA, Hattersley AT. Markers of β-Cell Failure Predict Poor Glycemic Response to GLP-1 Receptor Agonist Therapy in Type 2 Diabetes. Diabetes Care 2016;39:250

18. RetroMASTER - Retrospective Cohort MRC ABPI STratification and Extreme Response Mechanism in Diabetes [article online], 2015. Available from https://ClinicalTrials.gov/show/NCT02109978.

19. StartRight: Getting the Right Classification and Treatment From Diagnosis in Adults With Diabetes [article online], 2022. Available from https://ClinicalTrials.gov/show/NCT03737799.

20. Eason RJ, Thomas NJ, Hill AV, Knight BA, Carr A, Hattersley AT, McDonald TJ, Shields BM, Jones AG. Routine Islet Autoantibody Testing in Clinically Diagnosed Adult-Onset Type 1 Diabetes Can Help Identify Misclassification and the Possibility of Successful Insulin Cessation. Diabetes Care 2022;

21. Thomas NJ, Hill AV, Dayan CM, Oram RA, McDonald TJ, Shields BM, Jones AG, Group SS. Age of Diagnosis Does Not Alter the Presentation or Progression of Robustly Defined Adult-Onset Type 1 Diabetes. Diabetes Care 2023;

22. McDonald TJ, Colclough K, Brown R, Shields B, Shepherd M, Bingley P, Williams A, Hattersley AT, Ellard S. Islet autoantibodies can discriminate maturity-onset diabetes of the young (MODY) from Type 1 diabetes. Diabetic Medicine 2011;28:1028–1033

23. Wyatt RC, Grace SL, Brigatti C, Liberati D, Gillard BT, Marzinotto I, Shoemark D, Chandler MAM, Achenbach P, Piemonti L, The BOXSG, Long AE, Gillespie KM, Lampasona V, Williams AJK. Improved specificity of glutamate decarboxylase 65 autoantibody measurement using luciferase-based immunoprecipitation system (LIPS) assays. medRxiv 2023:2023.2007.2003.23292157

24. Achenbach PW, K.; Reiter, J.; Naserke, H.E.; Williams, A.J.K.; Bingley, P.J.; Bonifacio, E.; Ziegler, A-G. Stratificiation of Type 1 Diabetes Risk on the Basis of Islet Autoantibody Characteristics. Diabetes 2004;53:384–392

25. Curnock RM, Reed CR, Rokni S, Broadhurst JW, Bingley PJ, Williams AJK. Insulin autoantibody affinity measurement using a single concentration of unlabelled insulin competitor discriminates risk in relatives of patients with type 1 diabetes. Clin Exp Immunol 2012;167:67–72

26. Thomas NJ, Lynam AL, Hill AV, Weedon MN, Shields BM, Oram RA, McDonald TJ, Hattersley AT, Jones AG. Type 1 diabetes defined by severe insulin deficiency occurs after 30 years of age and is commonly treated as type 2 diabetes. Diabetologia 2019;62:1167–1172

27. Oram RA, Patel K, Hill A, Shields B, McDonald TJ, Jones A, Hattersley AT, Weedon MN. A Type 1 Diabetes Genetic Risk Score Can Aid Discrimination Between Type 1 and Type 2 Diabetes in Young Adults. Diabetes Care 2016;39:337

28. Bender C, Schlosser M, Christen U, Ziegler AG, Achenbach P. GAD autoantibody affinity in schoolchildren from the general population. Diabetologia 2014;57:1911–1918

29. Williams CL, Fareed R, Mortimer GLM, Aitken RJ, Wilson IV, George G, Gillespie KM, Williams AJK, Long AE. The longitudinal loss of islet autoantibody responses from diagnosis of type 1 diabetes occurs progressively over follow-up and is determined by low autoantibody titres, early-onset, and genetic variants. Clin Exp Immunol 2022;

30. Gu Y, Jia X, Vartak T, Miao D, Dong F, Jerram ST, Rewers M, Ferrara A, Lawrence JM, Yu L, Leslie RD, Leslie RD, Hawa MI, Pozzilli P, Beck-Nielsen H, Yderstraede K, Hunter S, Hadden D, Buzzetti R, Scherbaum W, Kolb H, Schloot NC, Seissler J, Schernthaner G, Tuomilehto J, Sarti C, De Leiva A, Brugues E, Mauricio D, Thivolet C, Lawrence JM, Ferrara A, Slezak JM, Quesenberry C, Saydah S, Yu L, Rewers M, the Action Lc, the Diabetes in Young Adults Study G. Improving clinical utility of GAD65 autoantibodies by electrochemiluminescence assay and clinical phenotype when identifying autoimmune adult-onset diabetes. Diabetologia 2021;64:2052-2060

31. Williams AJK, Lampasona V, Schlosser M, Mueller PW, Pittman DL, Winter WE, Akolkar B, Wyatt R, Brigatti C, Krause S, Achenbach P. Detection of Antibodies Directed to the N-Terminal Region of GAD Is Dependent on Assay Format and Contributes to Differences in the Specificity of GAD Autoantibody Assays for Type 1 Diabetes. Diabetes 2015;64:3239

32. Yu L, Miao D, Scrimgeour L, Johnson K, Rewers M, Eisenbarth GS. Distinguishing Persistent Insulin Autoantibodies With Differential Risk: Nonradioactive Bivalent Proinsulin/Insulin Autoantibody Assay. Diabetes 2011;61:179–186

33. Lampasona V, Pittman DL, Williams AJ, Achenbach P, Schlosser M, Akolkar B, Winter WE. Islet Autoantibody Standardization Program 2018 Workshop: Interlaboratory Comparison of Glutamic Acid Decarboxylase Autoantibody Assay Performance. Clin Chem 2019;65:1141–1152

34. Long AE, George G, Williams CL. Persistence of islet autoantibodies after diagnosis in type 1 diabetes. Diabetic Medicine 2021;38:e14712

35. Bingley PJ. Clinical applications of diabetes antibody testing. The Journal of clinical endocrinology and metabolism 2010;95:25–33

