## Supplemental Methods for "Autoantibodies to truncated GAD(96-585) antigen stratify risk of early insulin requirement in adult-onset diabetes"

ESM Supplemental Methods

Measurement of GADA IgG subclass response to full-length GAD65(1-585)

GADA IgG subclasses were determined using [35S]-methionine-labelled GAD65 and biotinylated IgG subclass-specific mouse anti-human monoclonal antibodies bound by Streptavidin-Sepharose beads (Sigma Aldrich, Dorset, UK). Mouse anti-human IgG subclass antibodies used were IgG1 (clone G17-1), IgG2 (clone G18-21), IgG3 (clone HP6047), and IgG4 (clone JDC-14). All were provided by BD Biosciences, SD, USA, other than IgG3 (clone HP6047) provided by Invitrogen, ThermoFisher, CA, USA). Non-specific binding was determined using a mouse anti-rat IgM monoclonal antibody (clone G53-238*) and all results were expressed in delta counts per minute (∆CPM) and converted to a SD score (SDS) (1). The SDS in the present study was based on 49 healthy schoolchildren; an SDS ≥3 was considered positive. To assess assay performance, a series of quality control (QC) measures were previously developed (C.L.W, Bristol, U.K.) and run in all assays alongside samples in the present study: 1) 50% suspension of Ethanolamine-blocked Protein G Sepharose in Tris buffered saline with Tween-20 buffer (TBST; 20 mM Tris, 150 mM NaCl, pH 7.4 + 0.5% Tween-20) was used as a total IgG positive QC, 2) pooled patient sera was used to develop two composite IgG subclass positive QCs (IgG1/IgG2 and IgG1/IgG3/IgG4; ≥3 SDS for present IgG subclass), and 3) sera from a healthy adult was used as a negative QC (<3 SDS).

Measurement of GADA affinity to full-length GAD65(1-585)

Briefly, serum (2µl in duplicate) was plated and incubated for 24 hours at 4°C with 25µl/well of [^125^I]-recombinant human GAD65 (1.88x10^-10^ mol/l; RSR Limited, Cardiff, U.K.) at five increasing quantities of unlabelled human GAD65 diluted in TBST (1.5x10^-15^, 1.5x10^-14^, 1.5x10^-13^, 1.5x10^-12^, and 1x10^-11^ mol/well) or TBST only. Immunocomplexes were precipitated for 1 hour (orbital shaking ~700rpm at 4°C) with 12.5µl/well PAS (PAS suspension washed four times in TBST buffer). After incubation, excess unbound [^125^I]-recombinant human GAD65 was excluded by centrifugation (503xg at 4°C for 3 mins) and five serial washes with TBST, transferred to microtubes (STARLABS, Milton Keynes, U.K.), and measured using a TopCount gamma counter (Perkin Elmer, Waltham, MA, USA); the results were expressed as mean CPM. IC_50_ and *K*_d_ values were calculated by non-linear regression analysis using a one-site model (*R^2^* >0.90), assuming equal antibody binding by labelled and unlabelled GAD65, on GraphPad Prism3 (GraphPad Software, San Diego, CA, USA) (2). The f-GADA affinity of each sample was expressed as the reciprocal *K*_d_ value (l/mol). Samples not fully competed (and therefore a non-linear regression was not fitted) were diluted 1:5 or 1:10 in TBST buffer and reanalysed for accurate f-GADA affinity assessment.

1.	Bonifacio E, Scirpoli M, Kredel K, Füchtenbusch M, Ziegler A-G. Early Autoantibody Responses in Prediabetes Are IgG1 Dominated and Suggest Antigen-Specific Regulation. The Journal of Immunology. 1999;163(1):525.

2.	Curnock RM, Reed CR, Rokni S, Broadhurst JW, Bingley PJ, Williams AJK. Insulin autoantibody affinity measurement using a single concentration of unlabelled insulin competitor discriminates risk in relatives of patients with type 1 diabetes. Clin Exp Immunol. 2012;167(1):67-72.
