## Supplemental Table for "Autoantibodies to truncated GAD(96-585) antigen stratify risk of early insulin requirement in adult-onset diabetes"

| Variable | f-GADA Positive T1D Reference Cohort | T2D (All) | T2D with f-GADA positivity | T2D with f-GADA negativity |
|---|---|---|---|---|
| **n** | 141 | 6,599 | 179 (2.7%) | 6,420 (97.3%) |
| **Male (%; 95% CI)** | 67 (48%; 39, 56) | 3,761 (57%; 56, 58) | 79 (56%; 48, 63) | 2,758 (57%; 56, 58) |
| **Ethnicity (%Caucasion; 95% CI)** | 136 (96%; 92, 99) | 6,597 (99.9%; 99.8, 99.9) | 177 (99%; 96.0, 99.9) | 6,420 (100%; 99, 100†) |
| **Age at Diagnosis (Years)** | 27.1 (24.4, 29.7) | 60.2 (60.0, 60.5) | 56.1 (54.1, 58.0) | 60.4 (60.1, 60.6) |
| **Duration of diabetes at latest follow up (years)** | 16.8 (14.5, 19.2) | 10.9 (10.7, 11.0) | 9.6 (8.7, 10.5) | 10.9 (10.8, 11.1) |
| **BMI (at first visit; kg/m$^2$)** | 25.3 (24.6, 26.0) | 31.8 (31.6, 31.9) | 29.8 (28.9, 30.8) | 31.8 (31.7, 32.0) |
| **Duration of diabetes at BMI (months)** | 17.6 (14.3, 21.0) | 44.0 (42.3, 45.8) | 50.4 (37.8, 63.0) | 43.9 (42.1, 45.6) |
| **f-GADA titre (WHO U/ml)** | 622 (488, 755) | 23.4 (19.2, 27.6) | 667 (546, 788) | 5.5 (5.4, 5.5) |
| **Duration at f-GADA (years)** | 18.0 (15.7, 20.4) | 6.8 (6.6, 6.9) | 6.1 (5.3, 7.0) | 6.8 (6.7, 6.9) |
| **Insulin treated within 5 years (%, 95% CI)** | 141 (100%; 100, 100) | 492 (7.5%; 6.9, 8.2) | 63 (35%; 28, 43) | 429 (6.7%, 6.1, 7.3) |
| **HbA1c (mmol/mol)*** | 72.2 (68.7, 75.6) | 63.3 (62.8, 63.8) | 66.6 (63.3, 69.9) | 63.2 (62.7, 63.7) |
| **HbA1c (%)*** | 8.8 (8.4, 9.1) | 7.9 (7.9, 8.0) | 8.3 (7.9, 8.6) | 7.9 (7.9, 8.0) |
| **Duration at HbA1c (years)** | 17.2 (13.9, 20.6) | 5.1 (5.0, 5.3) | 5.7 (4.5, 6.8) | 5.1 (5.0, 5.3) |
| **T1D GRS** | 0.274 (0.269, 0.279) | 0.228 (0.227, 0.229) | 0.243 (0.237, 0.249) | 0.228 (0.227, 0.229) |
| **C-peptide (pmol/l)** | 54.2 (29.4, 78.9) | 2312 (2228, 2396) | 1361 (1038, 1684) | 2369 (2283, 2454) |

Supplemental Table 1: Overall cohort characteristics. Data displayed as n(%; 95% CI) or mean (95% CI). *At latest follow-up. †One-sided 97.5% CI. T1D; type 1 diabetes. T2D; type 2 diabetes.

| Variable | DARE | GoDARTS | MRC Progressors | PRIBA | StartRight |
|---|---|---|---|---|---|
| **n** | 1,906 | 3,893 | 212 | 558 | 30 |
| **Male (%)** | 1,148 (60%; 58, 62) | 2,134 (55%(; 53, 56) | 133 (63%; 56, 70) | 326 (58%; 54, 63) | 20 (67%; 47, 83) |
| **Ethnicity (%Caucasion)** | 1,906 (100%; 100, 100*) | 3,893 (100%; 100, 100*) | 212 (100%; 98, 100*) | 558 (100%; 99, 100*) | 28 (93%; 78, 99) |
| **Age at Diagnosis (Years)** | 61.1 (61.0, 61.7) | 61.4 (61.0, 61.7) | 56.5 (55.3, 57.6) | 51.7 (50.9, 52.4) | 41.1 (36.7, 45.5) |
| **Duration of diabetes at latest follow up (years)** | 7.7 (7.4, 8.0) | 12.5 (12.4, 12.6) | 15.1 (14.4, 15.8) | 9.3 (8.8, 9.8) | 2.4 (2.2, 2.6) |
| **f-GADA titre (WHO U/ml)** | 23.8 (15.7, 31.8) | 17.2 (12.9, 21.5) | 21.6 (-0.99, 44.2) | 8.2 (8.0, 8.5) | 1108 (795, 1421) |
| **Duration of diabetes at f-GADA assessment (years)** | 7.9 (7.6, 8.2) | 5.5 (5.4, 5.6) | 14.1 (13.4, 14.8) | 9.2 (8.7, 9.7) | 0.49 (0.39, 0.59) |
| **f-GADA Positive (& sera available for further analysis) (%)** | 54 (2.8%; 2.1, 3.7) | 84 (2.2%; 1.7, 2.7) | 11 (5.2%; 2.6, 9) | 0 (0%; 0, 0.7*) | 30 (100%; 88, 100*) |

Supplemental Table 2: Overall type 2 diabetes cohort characteristics split by study. Data displayed as n(%; 95% CI) or mean (95% CI). *One-sided, 97.5% CI

| Variable | DARE | GoDARTS | MRC Progressors | StartRight |
|---|---|---|---|---|
| **n** | 54 | 84 | 11 | 30 |
| **Male (%; 95% CI)** | 32 (59%; 45, 72) | 40 (48%; 37, 59) | 8 (73%; 39, 94) | 20 (67%; 47, 83) |
| **Ethnicity (%Caucasion; 95% CI)** | 54 (100%; 93, 100*) | 84 (100%; 96, 100*) | 11 (100%; 72, 100*) | 28 (93%; 78, 99) |
| **Age at Diagnosis (Years)** | 56.5 (53.3, 59.8) | 61.5 (59.2, 63.9) | 53.3 (49.7, 56.9) | 41.1 (36.7, 45.5) |
| **Duration of diabetes at latest follow up (years)** | 8.9 (6.8, 10.9) | 12.2 (11.4, 13.0) | 12.7 (11.0, 14.4) | 2.4 (2.2, 2.6) |
| **f-GADA titre (WHO U/ml)** | 662 (429, 895) | 558 (393, 722) | 326 (-139, 793) | 1108 (765, 1421) |
| **Duration of diabetes at f-GADA assessment (years)** | 9.0 (6.9, 11.0) | 5.5 (4.8, 6.3) | 11.7 (10.0, 13.4) | 0.49 (0.39, 0.59) |

Supplemental Table 3: f-GADA positive type 2 diabetes cohort split by study. Data displayed as n(%; 95% CI) or mean (95% CI). *One-sided, 97.5% CI

| | Unadjusted Model | | Adjusted Model | |
|---|---|---|---|---|
| **Overall survival with f-GADA negatives** | **HR(95% CI)** | ***p* value** | **HR(95% CI)** | ***p* value** |
| **f-GADA Negative (reference)** | 1 | | 1 | |
| **f-GADA Positive, t-GADA Negative (vs. reference)** | 0.93 (0.23, 3.72) | 0.916 | 0.98 (0.24, 3.95) | 0.978 |
| **f-GADA Positive, t-GADA Positive (vs. reference)** | 8.40 (6.43, 10.99) | <0.001 | 7.06 (4.96, 10.06) | <0.001 |
| **Age of Diagnosis (per 1 year increase)** | | | 0.96 (0.95, 0.96) | <0.001 |
| **f-GADA Titre (per 100 unit increase)** | | | 1.00 (0.98, 1.03) | 0.744 |
| **Duration of Diabetes at f-GADA testing (per 1 year increase)** | | | 0.98 (0.97, 1.00) | 0.03 |

Supplemental Table 4: Hazard Ratios from Cox proportional regression model (unadjusted and adjusted) for time to insulin censored at 5 years (f-GADA and t-GADA positivity). f-GADA; full length GAD(1-585) autoantibody. t-GADA; truncated GAD(96-585) autoantibody.

| f-GADA IgG Isotype | T2D without early insulin requirement | T2D with early insulin requirement | T2D without vs. with early insulin requirement *p* value | T1D Reference Cohort | T1D RC vs T2D with early insulin requirement *p* value | T1D RC vs T2D without early insulin requirement *p* value |
|---|---|---|---|---|---|---|
| **IgG1** | 66 (87%; 77, 94) | 52 (96%; 87, 99.5) | 0.066 | 72 (95%; 87, 99) | 0.676 | 0.092 |
| **IgG2** | 20 (26%; 17, 38) | 10 (19%; 9.3, 31) | 0.298 | 14 (18%; 10, 29) | 0.989 | 0.243 |
| **IgG3** | 39 (51%; 40, 63) | 26 (48%; 34, 62) | 0.722 | 36 (47%; 36, 59) | 0.930 | 0.626 |
| **IgG4** | 15 (20%; 11, 30) | 7 (13%; 5.4, 25) | 0.310 | 16 (21%; 13, 32) | 0.234 | 0.840 |
| **IgG Isotype Combination** | | | | | | |
| **IgG1 Restricted** | 27 (39%; 28, 52) | 22 (42%; 29, 57) | 0.724 | 29 (38%; 27, 50) | 0.766 | 0.737 |

Supplemental Table 5: Individual IgG Subclass prevalence. Data shown as n(%; 95% CI). RC; Reference cohort. f-GADA; full length GAD(1-585) autoantibody. T1D; Type 1 Diabetes. T2D; Type 2 Diabetes.

| Characteristic | T2D IgG1-Restricted | T2D IgG-Unrestricted | IgG1-Restricted vs. IgG-Unrestricted *p* value | T1D Reference cohort | T2D f-GADA Negative |
|---|---|---|---|---|---|
| **n** | 53 (41%) | 76 (59%) | | 141 | 6,420 |
| **Age at Diagnosis (Years)** | 56.2 (52.4, 60.0) | 54.2 (51.0, 57.4) | 0.422 | 27.1 (24.4, 29.7) | 60.4 (60.1, 60.6) |
| **f-GADA Titre (WHO U/ml)** | 468 (283, 652) | 1130 (918, 1342) | p<0.0001 | 622 (488, 755) | 5.5 (5.4, 5.5) |
| **T1D Genetic Risk Score** | 0.248 (0.238, 0.258) | 0.248 (0.238, 0.257) | 0.955 | 0.274 (0.269, 0.279) | 0.228 (0.227, 0.229) |
| **C-Peptide (pmol/L)** | 1160 (706, 1614) | 1184 (814, 1555) | 0.931 | 54.2 (29.4, 78.9) | 2369 (2283, 2454) |
| **IA-2A Positive (%)** | 9 (18%; 8.8, 32) | 22 (31%; 20, 43) | 0.132 | 70 (50%; 41, 58) | 15 (0.6%; 0.3, 0.9)* |
| **ZnT8A Positive (%)** | 9 (18%; 8.8 32) | 18 (25%; 16, 37) | 0.390 | 49 (39%; 30, 48) | 28 (1.7%; 1.2, 2.5)[†] |
| **Insulin treated within 5 years (%)** | 22 (45%; 31, 60) | 30 (42%; 30, 54) | 0.724 | 141 (100%; 100, 100) | 429 (6.7%; 6.1, 7.3) |

Supplemental Table 6: Diabetes characteristics comparison between those with an IgG1-restricted and an IgG-unrestricted response in those f-GADA positive. Data displayed as n(%; 95% CI) or mean (95% CI) *n=2,607 tested for IA-2A. [†]Out of 1,615 tested for ZnT8A. f-GADA; full length GAD(1-585) autoantibody. T1D; Type 1 Diabetes. T2D; Type 2 Diabetes. IA-2A; islet antigen-2 autoantibody. ZnT8A; zinc transporter 8 autoantibody.

| | Unadjusted Model | | Adjusted Model | |
|---|---|---|---|---|
| **Survival analysis with f-GADA negatives** | **HR(95% CI)** | ***p* value** | **HR(95% CI)** | ***p* value** |
| **f-GADA Negative (reference)** | 1 | | 1 | |
| **f-GADA Positive, IgG1-Restricted (vs. reference)** | 8.43 (53.82, 12.21) | <0.001 | 6.74 (3.93, 11.54) | <0.001 |
| **f-GADA Positive, IgG-Unrestricted (vs. reference)** | 9.28 (6.04, 14.24) | <0.001 | 8.91 (5.61, 14.14) | <0.001 |
| **Age of Diagnosis (per 1 year increase)** | | | 0.96 (0.95, 0.97) | <0.001 |
| **f-GADA Titre (per 100 unit increase)** | | | 1.01 (0.97, 1.04) | 0.761 |
| **Duration of Diabetes at f-GADA testing (per 1 year increase)** | | | 0.99 (0.97, 1.00) | 0.082 |
| **Survival analysis IgG1-Restricted vs. IgG-Unrestricted (in those f-GADA positive)** | **Unadjusted Model** | | **Adjusted Model** | |
| **IgG-Unrestricted (reference)** | 1 | | 1 | |
| **IgG1-Restricted (vs. reference)** | 1.07 (0.62, 1.85) | 0.813 | 1.10 (0.60, 2.00) | 0.767 |
| **Age of Diagnosis (per 1 year increase)** | | | 0.94 (0.92, 0.96) | <0.001 |
| **f-GADA Titre (per 100 unit increase)** | | | 1.01 (0.98, 1.05) | 0.526 |
| **Duration of Diabetes at f-GADA testing (per 1 year increase)** | | | 0.89 (0.83, 0.95) | <0.001 |

Supplemental Table 7: Hazard Ratios from Cox proportional regression model (unadjusted and adjusted) for time to insulin censored at 5 years (IgG subclass response). f-GADA; full length GAD(1-585) autoantibody.

| | Unadjusted Model | |
|---|---|---|
| | **HR (95% CI)** | ***p* value** |
| **Respective Subclass not present** | 1 | |
| **IgG Subclass present** | | |
| **IgG1** | 3.0 (0.7, 12.1) | 0.133 |
| **IgG2** | 0.7 (0.4, 1.4) | 0.31 |
| **IgG3** | 0.9 (0.6, 1.6) | 0.845 |
| **IgG4** | 0.7 (0.3, 1.5) | 0.357 |

Supplemental Table 8: Hazard Ratios from Cox proportional regression model (unadjusted) for time to insulin censored at 5 years (IgG subclass present).

| | T2D with High affinity f-GADA | T2D with Moderate/low affinity f-GADA | T2D High vs. Moderate/low Affinity *p* value | T1D Reference cohort | T2D with f-GADA negativity |
|---|---|---|---|---|---|
| **n** | 114 (71%) | 47 (29%) | | 141 | 6,420 |
| **Age at Diagnosis (Years)** | 55.3 (52.9, 57.8) | 55.7 (51.8, 59.7) | 0.858 | 27.1 (24.4, 29.7) | 60.4 (60.1, 60.6) |
| **f-GADA Titre (WHO U/ml)** | 546 (409, 683) | 1167 (902, 1432) | <0.0001 | 622 (488, 755) | 5.5 (5.4, 5.5) |
| **T1D Genetic Risk Score** | 0.249 (0.242, 0.256) | 0.232 (0.219, 0.244) | 0.013 | 0.274 (0.269, 0.279) | 0.228 (0.227, 0.229) |
| **C-Peptide (pmol/L)** | 1195 (928, 1462) | 1247 (652, 1842) | 0.851 | 54.2 (29.4, 78.9) | 2369 (2283, 2454) |
| **IA-2A Positive (%)** | 21 (18%; 12, 27) | 13 (28%; 16, 43) | 0.192 | 70 (50%; 41, 58) | 15 (0.6%; 0.3, 0.9)* |
| **ZnT8A Positive (%)** | 17 (15%; 8.9, 23) | 13 (28%; 16, 43) | 0.059 | 49 (39%; 30, 48) | 28 (1.7%; 1.2, 2.5)† |
| **Insulin treated within 5 years (%)** | 45 (39%; 30, 49) | 16 (34%; 21, 49) | 0.518 | 141 (100%; 100, 100) | 429 (6.7%; 6.1, 7.3) |

Supplemental Table 9: Diabetes characteristics comparison between those with higher and lower affinity f-GADA. Data displayed as n(%; 95% CI) or mean (95% CI). *Out of 2,607 tested. †Out of 1,615 tested for ZnT8A. T1D; Type 1 Diabetes. T2D; Type 2 Diabetes. f-GADA; full length GAD(1-585) autoantibody. IA-2A; islet antigen-2 autoantibody. ZnT8A; zinc transporter 8 autoantibody.

| | Unadjusted Model | | Adjusted Model | |
|---|---|---|---|---|
| **Survival with f-GADA negatives** | **HR(95% CI)** | ***p* value** | **HR(95% CI)** | ***p* value** |
| **f-GADA Negative (reference)** | 1 | | 1 | |
| **f-GADA Positive, Moderate/Low Affinity f-GADA (vs. reference)** | 6.53 (3.97, 10.77) | <0.001 | 4.67 (2.45, 8.89) | <0.001 |
| **f-GADA Positive, High Affinity f-GADA (vs. reference)** | 7.62 (5.60, 10.35) | <0.001 | 6.34 (4.40, 9.13) | <0.001 |
| **Age of Diagnosis (per 1 year increase)** | | | 0.957 (0.948, 0.965) | <0.001 |
| **f-GADA Titre (per 100 unit increase)** | | | 1.02 (0.984, 1.05) | 0.326 |
| **Duration of Diabetes at f-GADA testing (per 1 year increase)** | | | 0.983 (0.967, 0.999) | 0.034 |
| **High vs. Moderate/low Affinity in those f-GADA positive** | **Unadjusted Model** | | **Adjusted Model** | |
| **Moderate/Low Affinity f-GADA (reference)** | 1 | | 1 | |
| **High Affinity f-GADA (vs. reference)** | 1.13 (0.64, 2.01) | 0.664 | 1.17 (0.63, 2.17) | 0.621 |
| **Age of Diagnosis (per 1 year increase)** | | | 0.941 (0.922, 0.960) | <0.001 |
| **f-GADA Titre (per 100 unit increase)** | | | 1.02 (0.983, 1.05) | 0.348 |
| **Duration of Diabetes at f-GADA testing (per 1 year increase)** | | | 0.879 (0.827, 0.934) | <0.001 |

Supplemental Table 10: Hazard Ratios from Cox proportional regression model (unadjusted and adjusted) for time to insulin censored at 5 years (f-GADA affinity category). f-GADA; full length GAD(1-585) autoantibody.

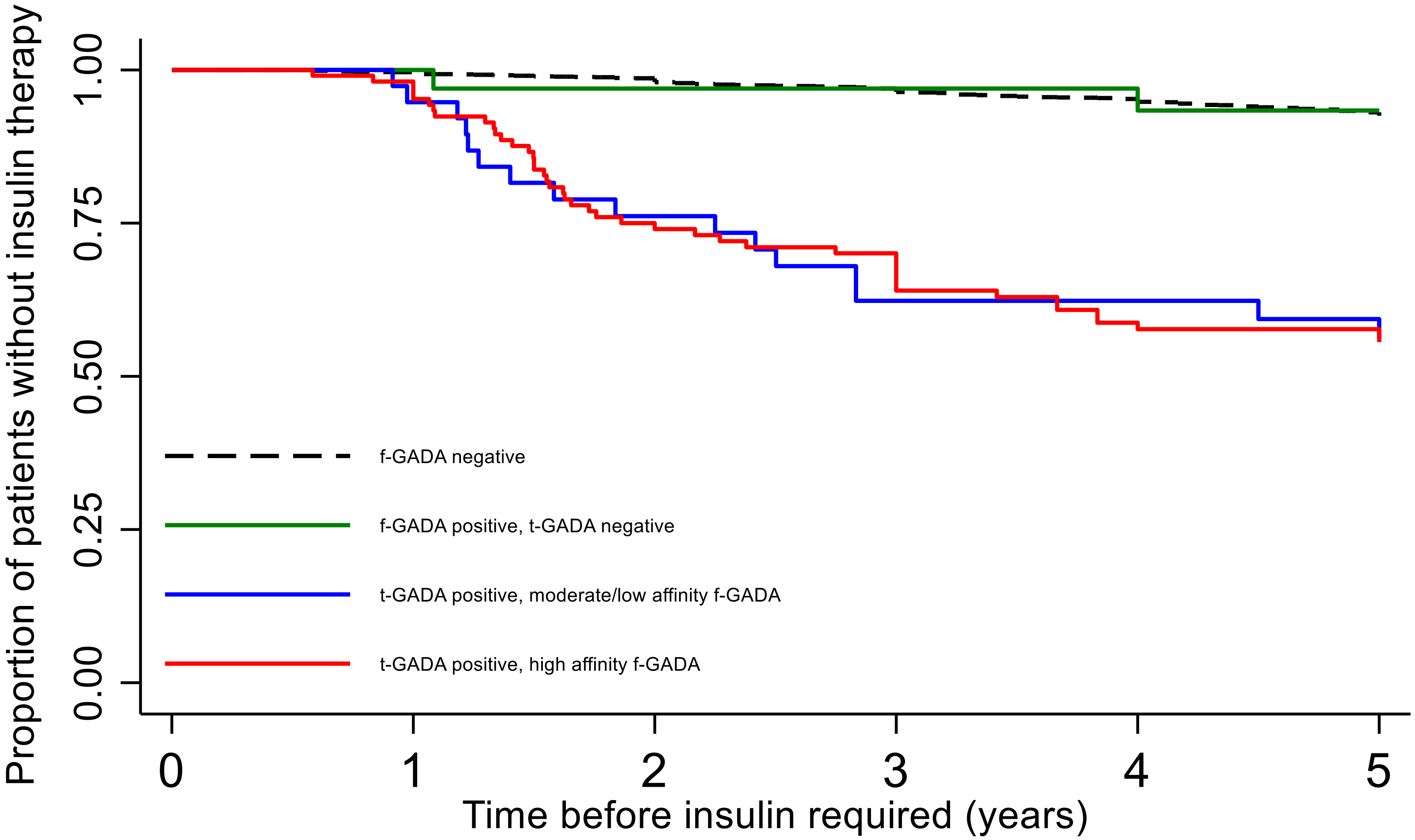

| Number at risk | 0 | 1 | 2 | 3 | 4 | 5 |
|---|---|---|---|---|---|---|
| f-GADA - | 6420 | 6327 | 5971 | 5563 | 5241 | 4930 |
| f-GADA +, t-GADA - | 34 | 34 | 28 | 28 | 27 | 25 |
| t-GADA +, Moderate/low affinity f-GADA | 39 | 36 | 28 | 22 | 21 | 19 |
| t-GADA +, High affinity f-GADA | 106 | 103 | 77 | 69 | 56 | 55 |

Supplemental Figure 1: Kaplan-Meier plots of probability of requiring insulin therapy during 5-year follow-up, in those clinically diagnosed with type 2 diabetes. Stratified by t-GADA positivity and f-GADA affinity. Solid lines indicate f-GADA positivity. Green line indicates t-GADA negative. Red line is t-GADA positive, high f-GADA affinity. Blue line indicates t-GADA positive, moderate/low f-GADA affinity
